# Cognitive Screening Tools for Dementia Detection in Primary Healthcare Centres in India: A Scoping Review

**DOI:** 10.1101/2024.12.04.24318472

**Authors:** R Jeevitha Gowda

**Author notes:** Corresponding author; Email ID.

## Abstract

**Background:** Dementia is a growing public health concern in India, with an increasing prevalence among the elderly population. Early detection is crucial for effective intervention. Primary healthcare (PHC) centres play a vital role in identifying cognitive impairment; however, the effectiveness of cognitive screening tools in these settings is questionable.

**Objective:** This scoping review explores the cognitive screening tools available for dementia detection in PHC centres in India, assesses their effectiveness, and identifies the need for their improvement and adaptation.

**Methods:** A systematic search was conducted in PubMed, Scopus, and Google Scholar for studies published up to October 2024. A total of 29 studies were identified, indicating that tools such as the Mini-Mental State Examination (MMSE) and the Montreal Cognitive Assessment (MoCA) are frequently used. However, these tools face significant challenges related to educational background and language comprehension, impacting their effectiveness.

**Conclusion:** There is an urgent need for culturally and linguistically appropriate cognitive screening tools in PHC settings in India to enhance the early detection of dementia.

## 1. Introduction

Dementia is a progressive neurodegenerative disorder characterised by a decline in cognitive function, impacting memory, thinking, and social abilities to the extent that it interferes with daily life ^1^. As the population ages, dementia has become a significant public health challenge globally, with an increasing prevalence observed in India. With over 140 million people aged 60 years and above, the country is facing a burgeoning dementia burden, necessitating effective screening strategies to facilitate early diagnosis and intervention ^2^.

Screening for dementia is crucial in identifying individuals who may be experiencing cognitive decline before the condition significantly impacts their quality of life. Early detection allows for timely interventions, support for patients and families, and the potential to slow the progression of symptoms^3^. Primary healthcare (PHC) centres serve as the first point of contact for individuals seeking medical assistance, making them ideal settings for cognitive screening. Given their accessibility, PHCs play a vital role in reaching vulnerable populations, especially in rural and underserved areas where specialized dementia services may be lacking ^4^.

In India, several cognitive screening tools have been implemented in PHC settings, including the Mini-Mental State Examination (MMSE) ^5^ and the Montreal Cognitive Assessment (MoCA) ^6^. While these tools aim to assist healthcare providers in identifying cognitive impairment, their effectiveness has been questioned due to factors such as educational background and language barriers among patients^7^. This review addresses the current landscape of cognitive screening tools in Indian PHCs, highlighting their effectiveness, associated challenges, and barriers to successful implementation. By providing a comprehensive overview, this review aims to identify gaps in the existing tools and underscore the need for culturally and linguistically appropriate alternatives tailored to the diverse population of India.

## 2. Research Question

The objective of this scoping review is to identify and assess the effectiveness of cognitive screening tools used for dementia screening in primary healthcare centres in India. The two review questions include: (1) What cognitive screening tools are utilized for dementia detection in primary healthcare centres (PHC) in India? and (2) how effective are these tools considering the educational and language barriers faced by the diverse Indian population?

## 3. Objective

The primary objectives of this scoping review are to:

1. Identify cognitive screening tools used for dementia detection in primary healthcare centres in India.
2. Assess the effectiveness of these tools, considering educational and language barriers.
3. Highlight the need for culturally and linguistically appropriate cognitive screening tools in PHC settings.

## 4. Methods

### 4.1. Study Design

This scoping review was conducted and reported following the Preferred Reporting Items for Systematic Reviews Meta-Analyses guidelines for scoping review and the steps of PRISMA 2018 ^8^

### 4.2. Protocol and Registration

The protocol for this scoping review was registered with the Open Science Framework (OSF) https://osf.io/58jvw/

### 4.3. Ethics Approval

No ethical approval was required, as this review is based on published literature.

### 4.4. Eligibility criteria

Eligibility criteria were based on the Participant, Concept, Context (PCC) framework to ensure alignment with the study objectives. Studies meeting the following criteria were included:

Participants: Studies examining patients screened for dementia or healthcare providers administering cognitive screening tools in primary healthcare (PHC) settings in India.

Concept: Studies focusing on the use, effectiveness, or validation of cognitive screening tools for dementia detection, particularly in PHC settings.

Context: Research conducted in PHC settings in India, addresses challenges such as literacy levels, language barriers, and cultural diversity.

Types of Sources: Peer-reviewed articles, conference proceedings, and grey literature published in English before October 2024. Articles without full-text access, conference abstracts, and non-research publications (e.g., editorials, commentaries), articles published before 1995 were excluded.

### 4.5. Data Search Strategy

A systematic search was conducted in multiple databases using keywords related to cognitive screening tools and dementia detection in PHC settings in India including Boolean operators (Tabe l). The search strategy included terms such as “cognitive screening,” “dementia,” “primary healthcare,” and “India.” Studies published in English up to October 2024 were included. Relevant articles from conferences and grey literature were also included using Google. The reference list of the selected articles was also checked for relevant studies. The complete search process led to a selection of 29.

The search encompassed the following databases: Tabel 1

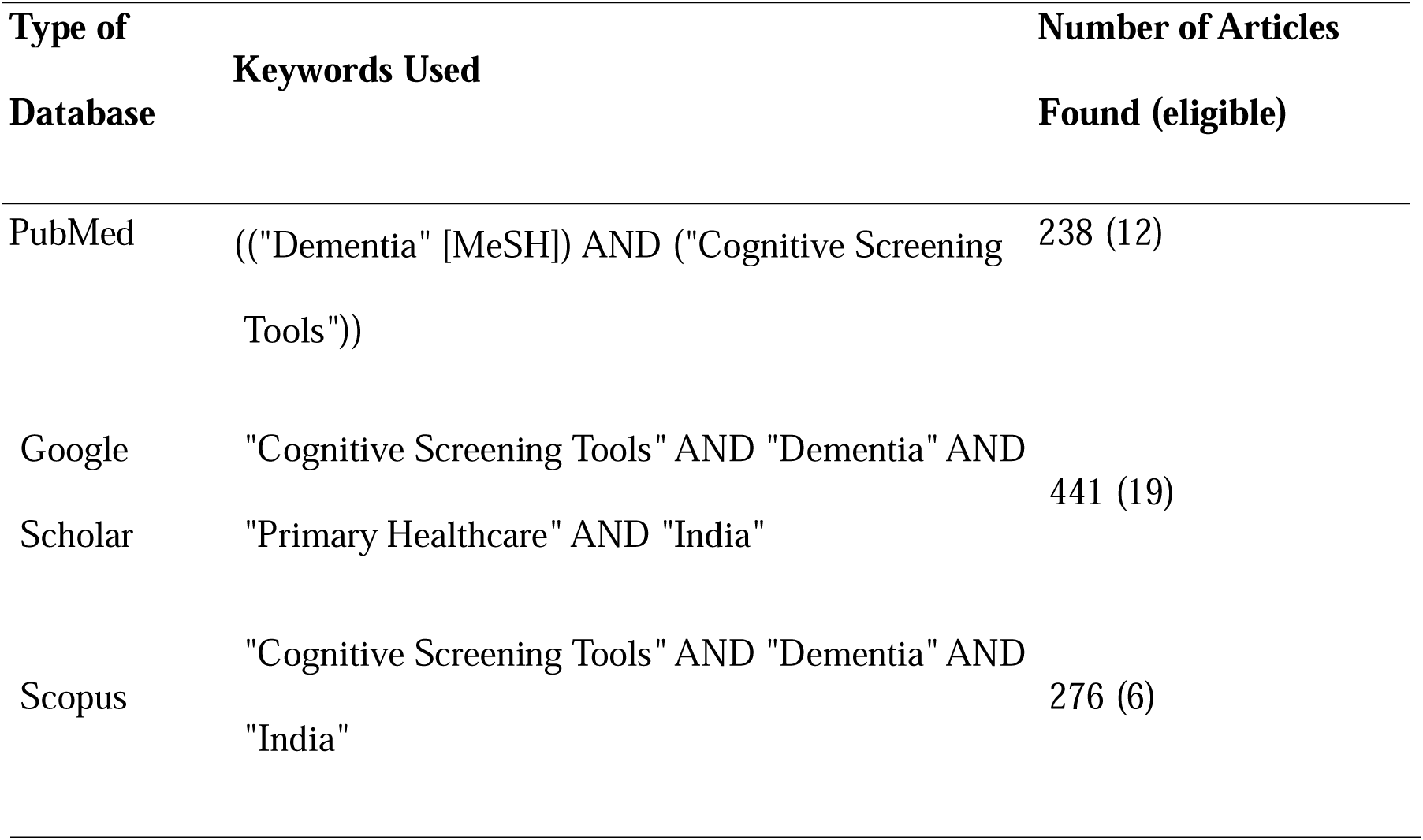

### 4.6. Study Selection Criteria

Inclusion criteria for selecting studies included:

- Studies focusing on cognitive screening tools for dementia detection.
- Research conducted in primary healthcare settings in India.
- Articles published in English.

Exclusion criteria included:

- Studies not focused on dementia or cognitive screening tools.
- Non-primary healthcare settings.
- Articles published before 1995

The selection process followed a structured methodology. Titles and abstracts of articles were reviewed for relevance by two independent (JR and PG) reviewers. Studies unrelated to cognitive screening tools or not conducted in PHC settings in India were excluded. Then full texts of potentially eligible articles were obtained and assessed against the inclusion criteria. Later, for the final selection discrepancies were resolved through discussion, and a final list of 29 studies was selected for inclusion in the scoping review. The steps of the screening process are illustrated in the PRISMA flow diagram in Figure 1.

**Figure 1:**
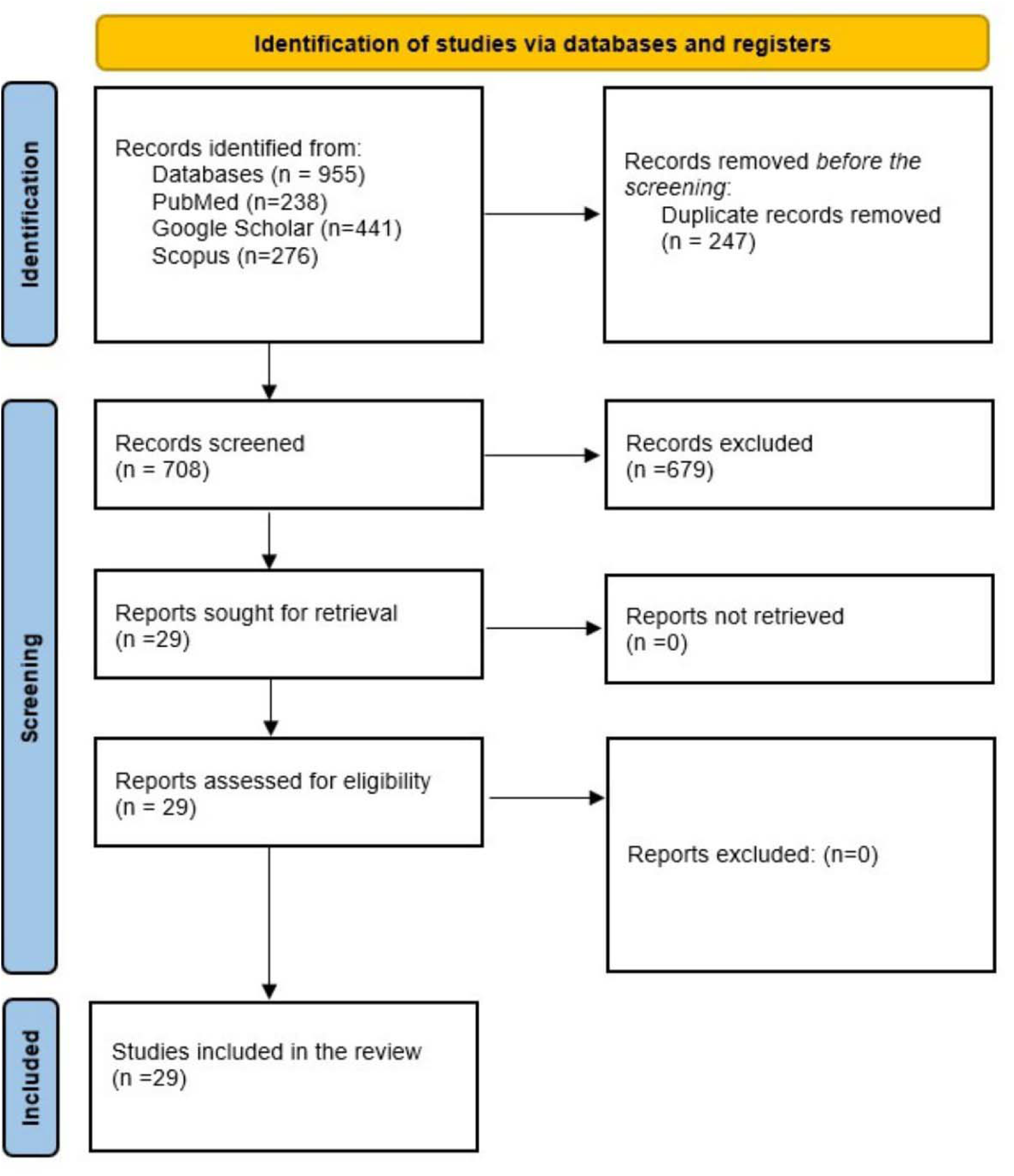
PRISMA flow diagram.

### 4.7. Data Extraction and presentation

Two independent reviewers (JR and PG) used a standardized data extraction form to gather key information from the included studies. Extracted data included, cognitive screening tools evaluated, reported barriers and facilitators for tool implementation, recommendations for tool adaptations in PHC settings and data Analysis and Presentation. Data were synthesized thematically to address the research questions. The findings were categorized as types of cognitive screening tools used in PHC settings, evaluation of tool performance, focusing on sensitivity, specificity, and applicability in low-literacy and linguistically diverse populations and limitations in implementation, including literacy, language, resource constraints, and cultural relevance.

## 5. Results

This scoping review identified various cognitive screening tools utilized in primary healthcare centres (PHC) for dementia detection in India. The analysis highlights the available tools, their effectiveness, associated issues, and barriers impacting their implementation. A total of 29 studies met the inclusion criteria, identifying several cognitive screening tools used in PHC settings.

### Study Selection

A total of 955 records were identified initially and 247 duplicates were removed. 679 records were excluded after reviewing titles and abstracts. 29 studies were selected after full-text review. The flow chart of the process is given in Figure 1.

### Study Characteristics

#### 5.1. Cognitive Screening Tools Used in India’s PHCs

This section presents the cognitive screening tools used in India’s PHC settings, their effectiveness, and the challenges posed by educational and language barriers.

Cognitive screening tools are essential for the early detection of dementia and cognitive impairments. Several cognitive screening instruments in India are commonly employed in PHCs, each with unique strengths and limitations. These tools vary in their focus, administration time, and cultural applicability. The evaluation of various cognitive screening tools reveals a mixed landscape of effectiveness in detecting cognitive impairments within primary health care (PHC) settings.

Table 2 has the key characteristics of these cognitive screening tools, including their administration time, available languages, effectiveness in PHC settings, and notable limitations. This comparative overview will help highlight the strengths and weaknesses of each tool, guiding healthcare providers in selecting the most suitable assessments for their patient populations.

**Table 2:**
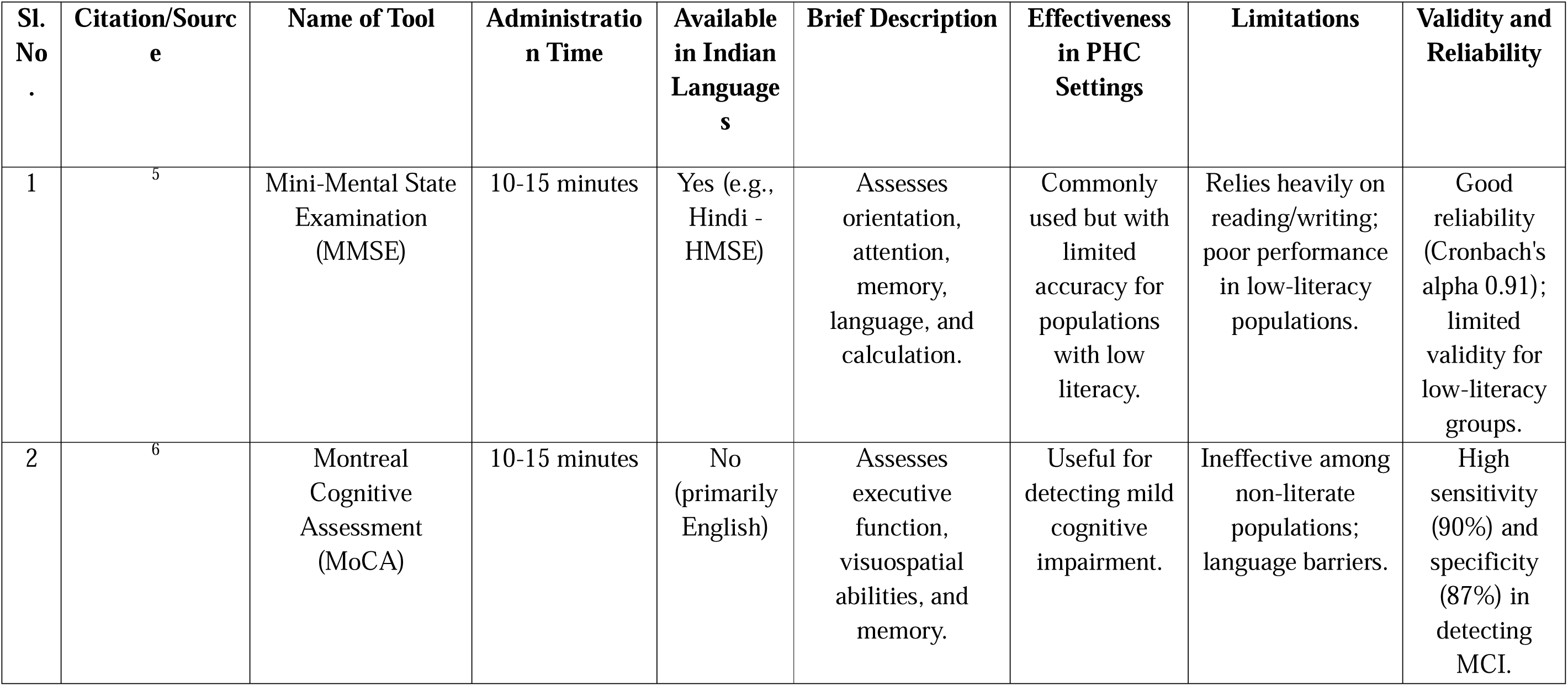
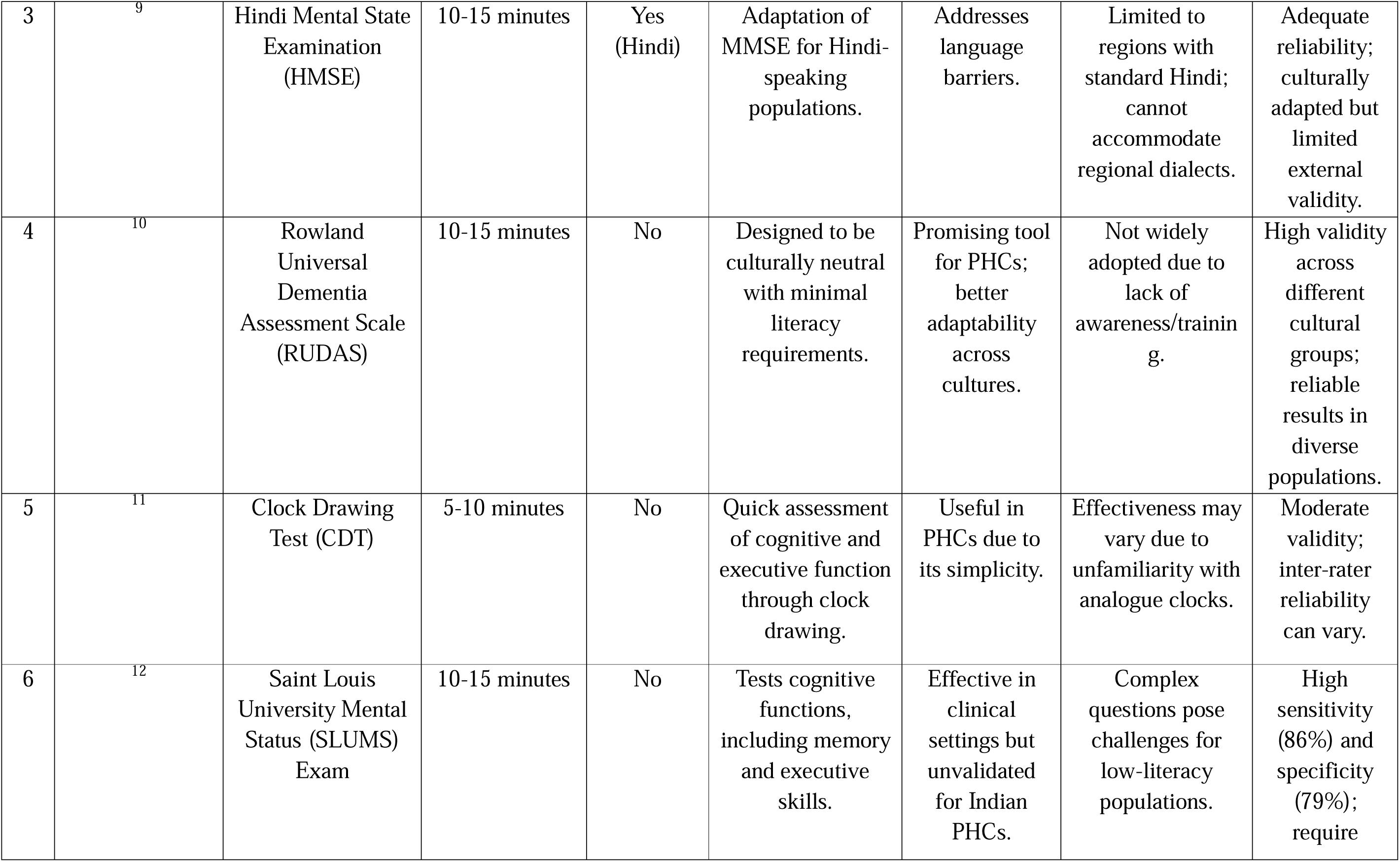
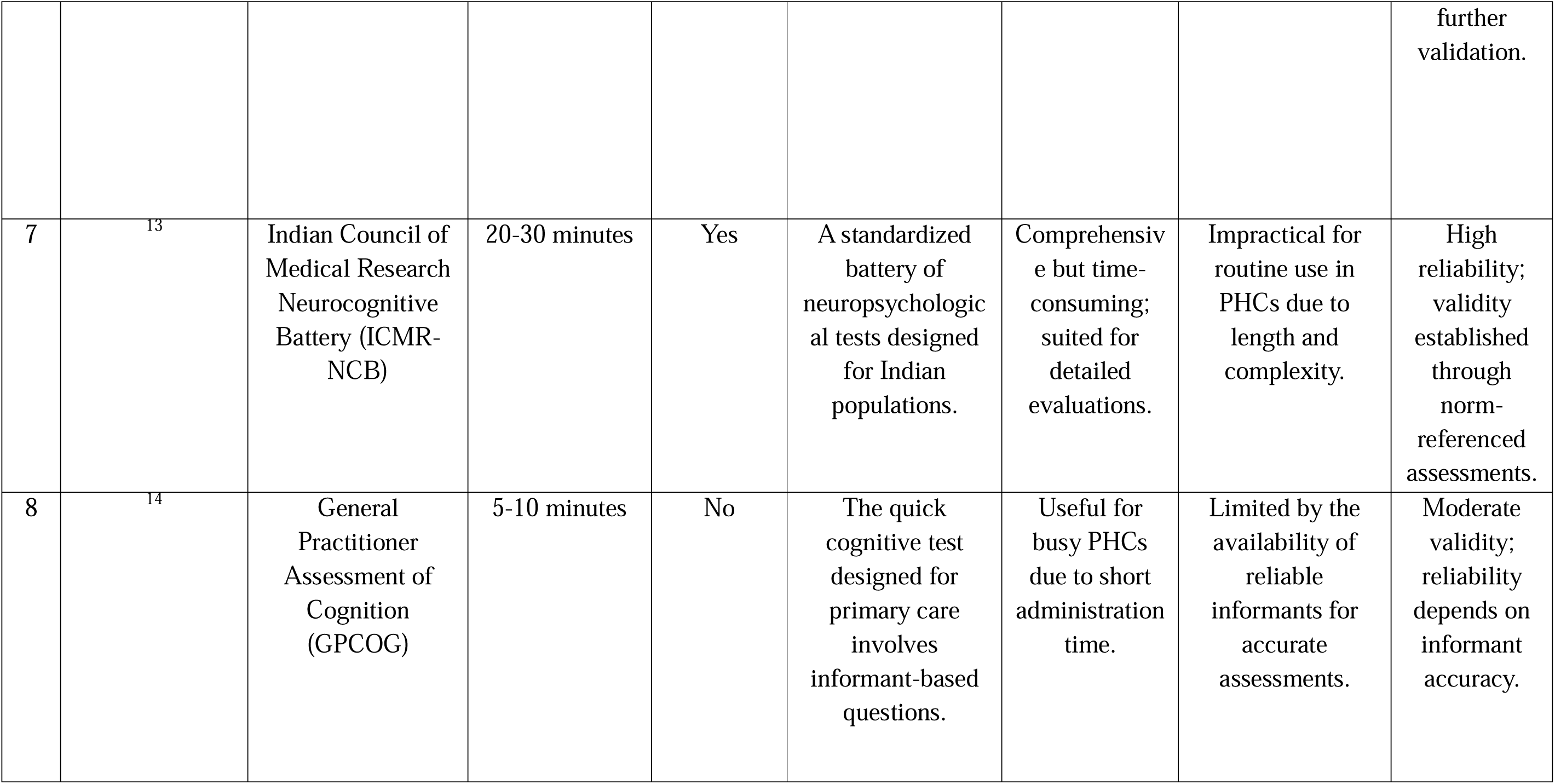
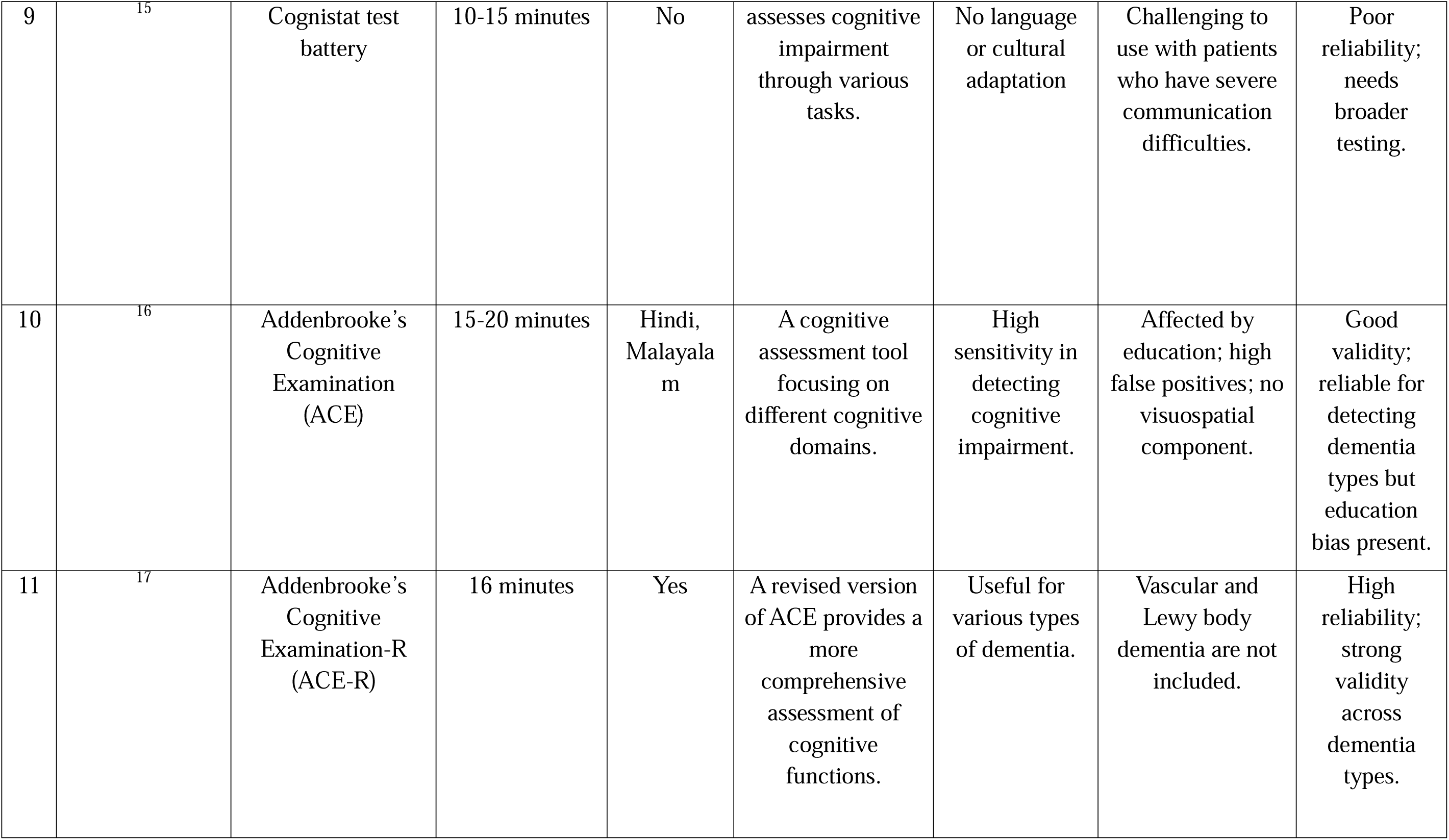
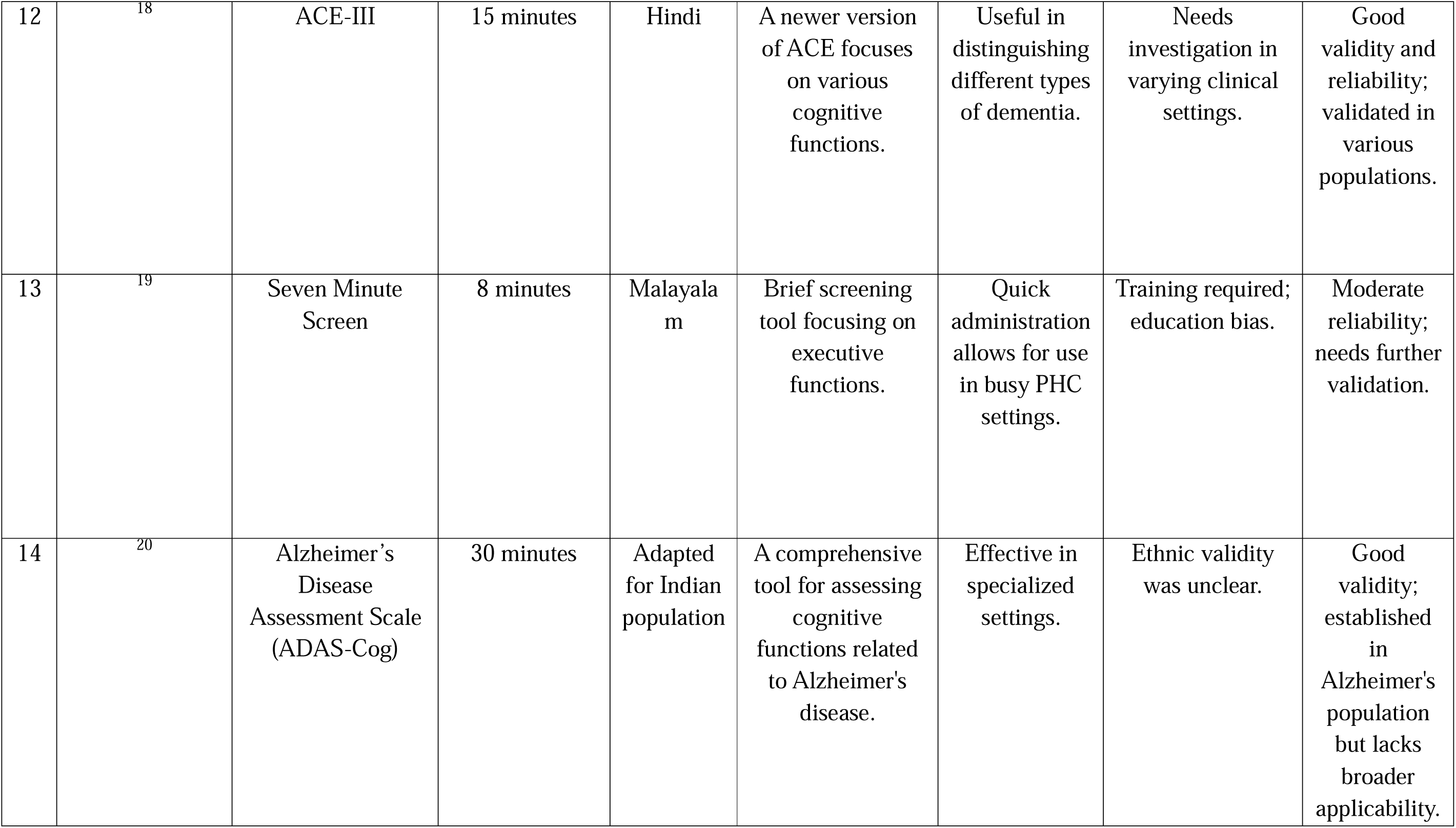
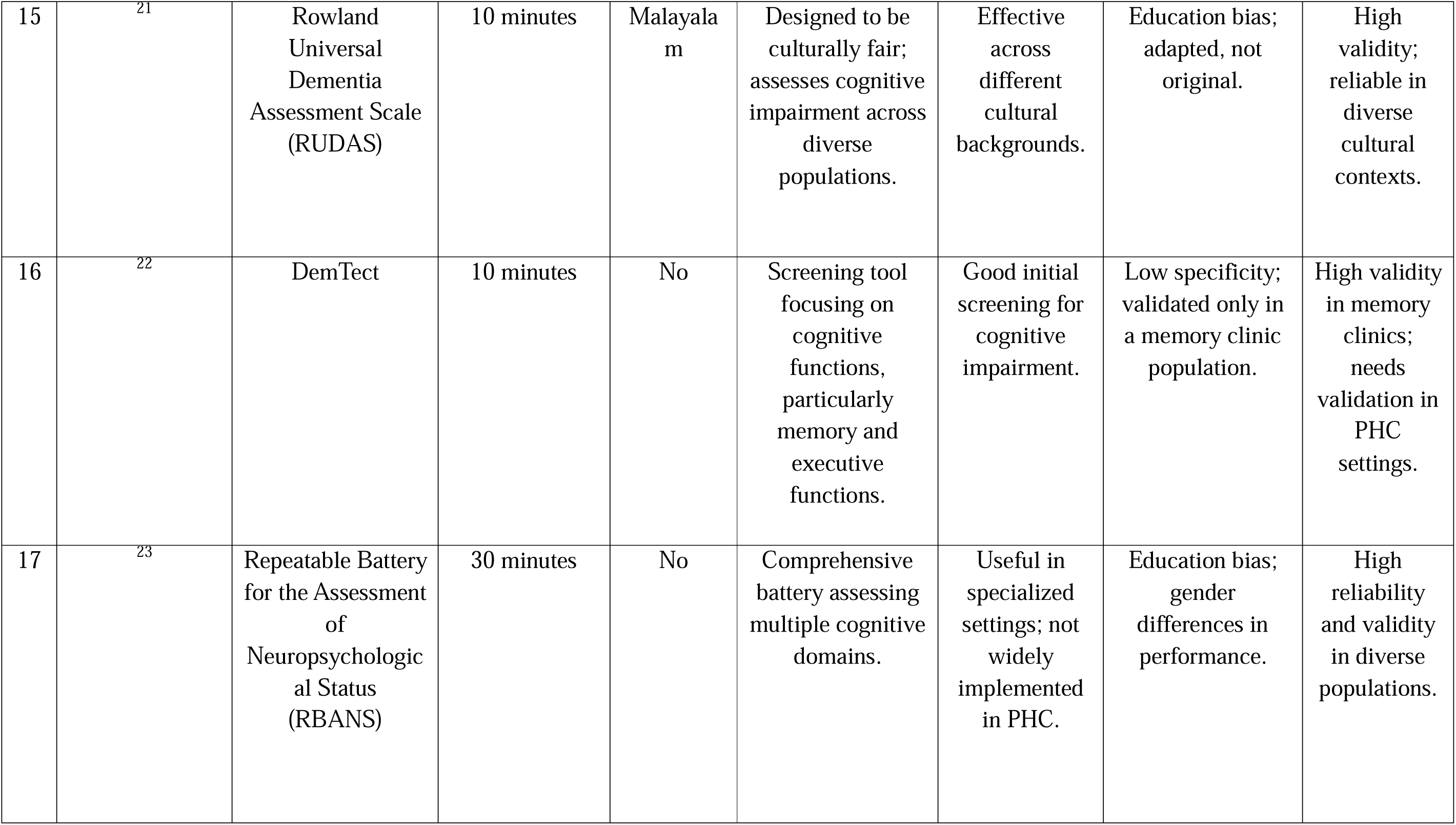
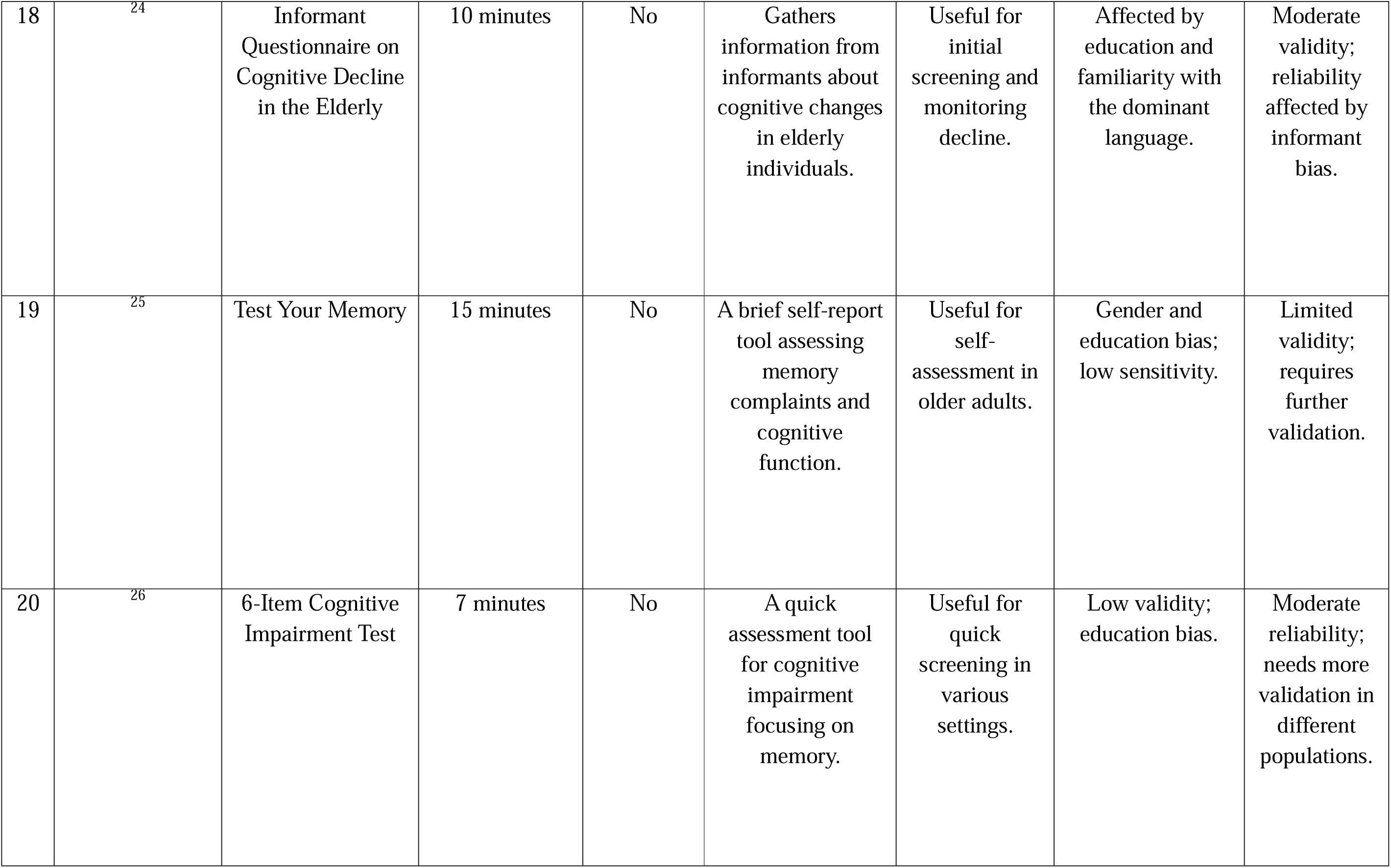
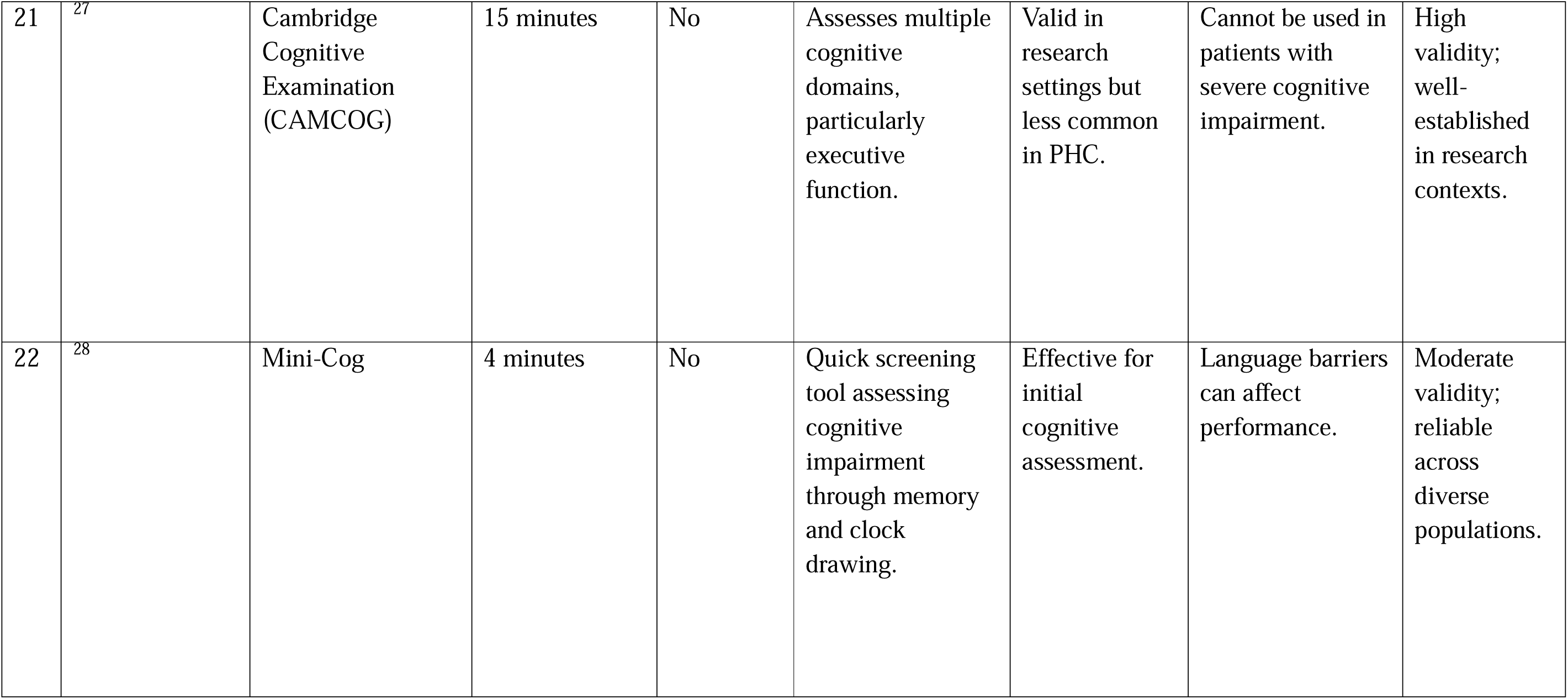
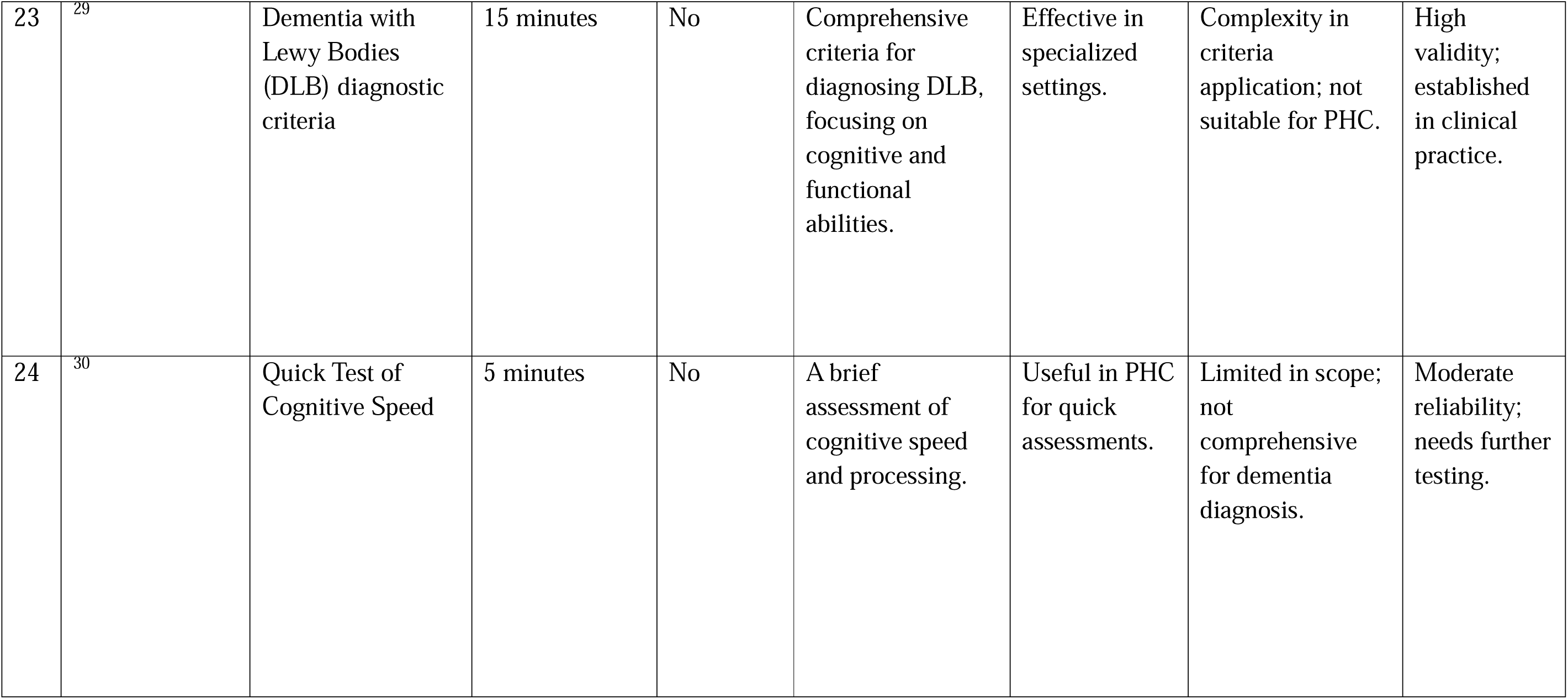
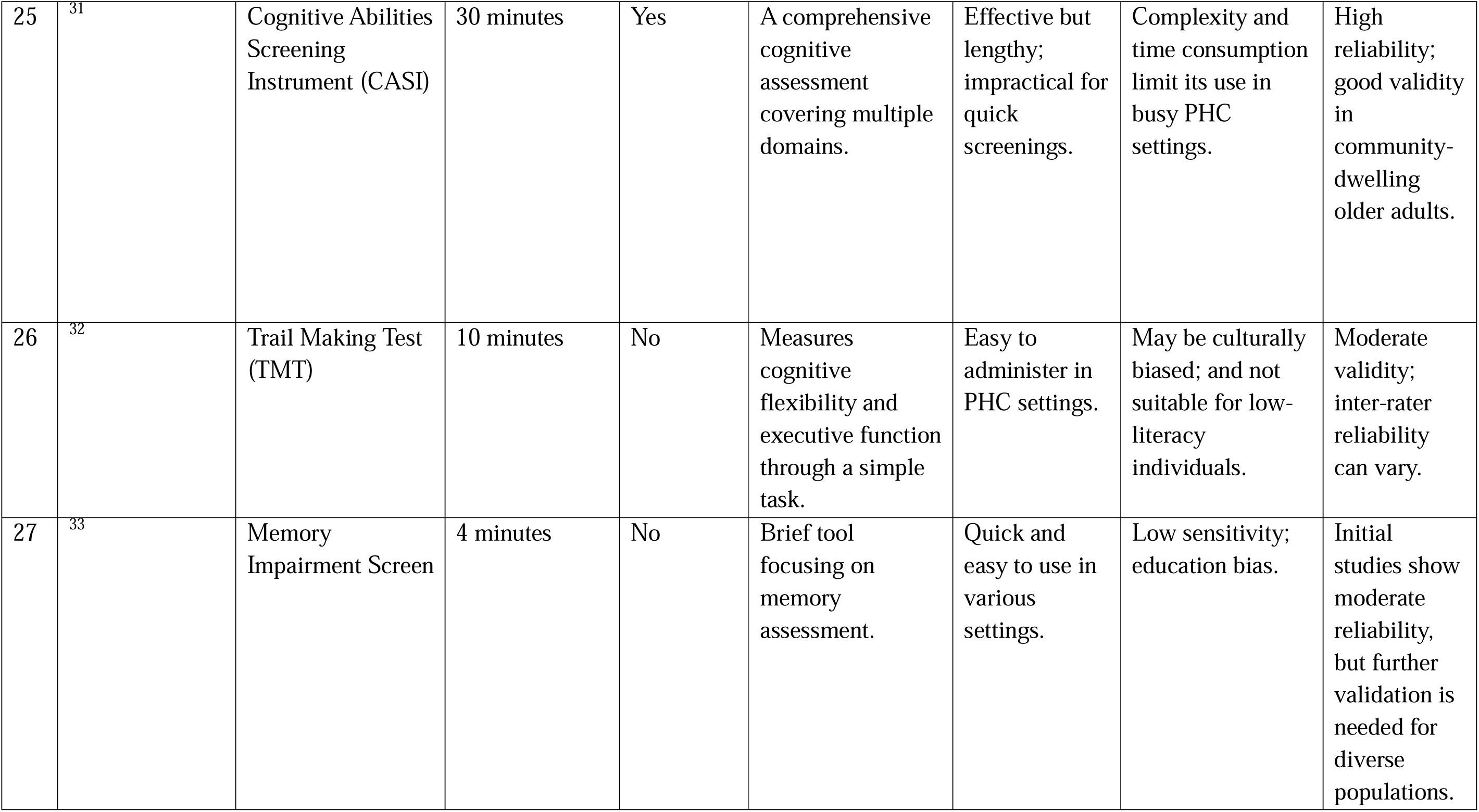
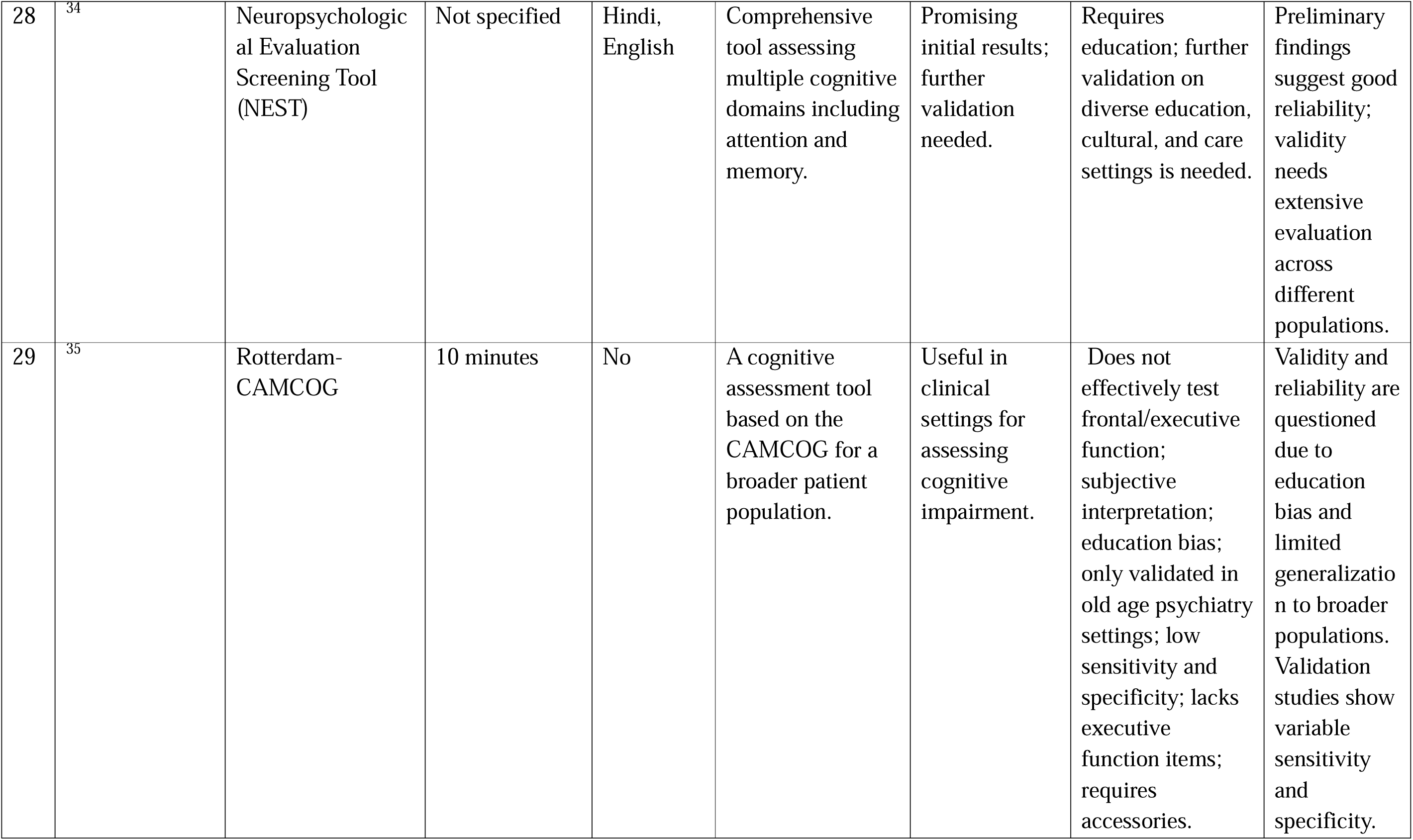
Cognitive screening tools with their key characteristics.

#### 5.2. Effectiveness of Cognitive Screening Tools in PHC Settings

The evaluation of various cognitive screening tools reveals a mixed landscape of effectiveness in detecting cognitive impairments within primary health care (PHC) settings. The analysis included 29 cognitive assessment instruments, highlighting their description, administration time, effectiveness, and limitations.

#### 1. Tool Availability and Language Adaptation

A significant number of tools, such as the Mini-Mental State Examination (MMSE) and the Hindi Mental State Examination (HMSE), are available in Indian languages, which facilitates their use in Hindi-speaking populations. The AIIMS Cognitive Screening Tool is also notable for its flexibility in language, catering to multiple Indian languages. However, many commonly used tools, including the Montreal Cognitive Assessment (MoCA) (Tavares-Júnior et al., 2019)and Addenbrooke’s Cognitive Examination (ACE-R), primarily exist in English, which can limit their accessibility and effectiveness among non-literate and low-literacy populations. Considering the diverse languages among the Indian population the current tools don’t overcome the language bias.

#### 2. Administration Time and Practicality

Most tools require approximately 10 to 15 minutes for administration, aligning well with the demands of busy PHC settings. Quick assessment tools, such as the General Practitioner Assessment of Cognition (GPCOG) and the Mini-Cog, are particularly advantageous, requiring only 5 to 10 minutes. In contrast, more comprehensive tools like the Indian Council of Medical Research Neurocognitive Battery (ICMR-NCB) and the AIIMS Neuropsychological Battery are lengthy (20-45 minutes), making them impractical for routine use in PHC due to the time constraints faced by healthcare providers.

#### 3. Effectiveness in Cognitive Impairment Detection

The effectiveness of these tools varies significantly. The MoCA and ACE-R are effective in identifying mild cognitive impairment (MCI), particularly in clinical environments, but face challenges in PHC settings due to their reliance on literacy and numeracy skills. Tools like the Rowland Universal Dementia Assessment Scale (RUDAS) show promise due to their culturally neutral approach and minimal literacy requirements, making them suitable for diverse populations.

The AIIMS Dementia Screening Questionnaire and the Addenbrooke’s Cognitive Examination (ACE) have demonstrated high sensitivity in detecting cognitive impairments, aiding in early diagnosis. However, the limitations of tools, such as the MMSE and MoCA, are evident, as they may not perform adequately among individuals with low educational backgrounds, potentially leading to both overdiagnosis and underdiagnosis.

#### 4. Challenges and Limitations

Common limitations across the tools include reliance on reading and writing, which affects their accuracy in populations with low literacy. Language barriers and the absence of culturally validated tools further complicate the screening process. Many tools have not been adequately tested in diverse cultural contexts, which raises concerns about their validity and reliability. Furthermore, the training required for administering certain tools can pose additional challenges, particularly in rural PHC settings where resources and trained personnel may be limited.

While many cognitive screening tools exhibit promising effectiveness for detecting cognitive impairments in PHC settings, significant challenges remain. The need for culturally and linguistically adaptable tools is critical to enhance their applicability. Addressing the limitations of existing tools through further research and validation studies will be essential for improving early detection and intervention strategies in the Indian population. Enhanced training for healthcare providers and the development of streamlined, user-friendly assessment tools will contribute to better cognitive health outcomes in primary care settings.

### 5.3. Barriers to Implementation

The implementation of cognitive screening tools in Primary Health Care (PHC) settings is significantly hindered by several barriers. Identifying and addressing these obstacles is crucial for improving the effectiveness of cognitive assessments, particularly in diverse and resource-constrained environments. This section explores the key barriers identified through the review.

#### 1. Educational Background

One of the most prominent barriers to effective cognitive screening is the educational background of patients. Individuals with lower levels of education often struggle to understand the tasks and questions posed by screening tools. This challenge can lead to poor performance that does not accurately reflect their true cognitive status ^36^. Many cognitive tools, including the MIS and NEST, require participants to follow tasks involving memory recall, attention, and verbal fluency. However, individuals with low literacy levels may struggle to complete these assessments accurately, leading to false negatives or overdiagnoses. This can significantly distort the assessment of cognitive abilities, resulting in improper care decisions. Also, tools like the Rotterdam-CAMCOG show education bias, performing poorly among participants with limited reading and numerical skills.

Future tools need to incorporate non-verbal components or simplified instructions to reduce educational bias. Consequently, the cognitive screening results may yield misleading interpretations, resulting in either overdiagnosis or underdiagnosis of cognitive impairments.

This mismatch between actual cognitive ability and test performance can adversely affect clinical decision-making and the provision of appropriate care ^37^.

#### 2. Language Barriers

Language barriers pose a significant challenge in the implementation of cognitive screening tools. The absence of culturally and linguistically appropriate versions of these tools can hinder effective communication during assessment. Many screening instruments primarily exist in English or a few widely spoken languages, limiting their accessibility for patients who are not proficient in these languages ^38^. The MoCA, MMSE and Rotterdam-CAMCOG are available only in a limited set of languages, making them unsuitable for use across India’s linguistically diverse regions. Further, they may not be sufficient for rural or tribal populations. Inappropriate language use can confuse patients, undermining their confidence in the healthcare system and reducing the reliability of cognitive assessments.

Language-appropriate versions of screening tools, validated for regional languages and cultural contexts, should be developed for more inclusive assessments. When patients cannot comprehend the questions or instructions, their ability to respond accurately is compromised, leading to false negatives or positives. This linguistic gap not only affects the reliability of the results but also undermines the patient’s confidence in the healthcare system.

### 5.4. Resource Constraints

Resource constraints within many PHC centres further complicate the effective implementation of cognitive screening tools. Many PHC centres in India face resource shortages, including a lack of trained personnel, space, and funding ^39^. Tools such as Rotterdam-CAMCOG, RUDAS, and AIIMS Neuro batteries require a kit, accessories and specific infrastructure, which are not always feasible in low-resource settings. Additionally, the absence of dedicated training programs makes it difficult for healthcare providers to integrate these tools into their routines.

Tools which aim to simplify administration should be further refined to function with minimal resources, ensuring they are usable in PHC environments. Also, the lack of resources can lead to inadequate training opportunities for healthcare providers, preventing them from acquiring the skills needed to administer these tools effectively ^40^. As a result, even if cognitive screening tools are available, the lack of trained personnel may limit their practical use in everyday clinical practice.

### 5.5. Provider Training Needs

A critical barrier to the effective implementation of cognitive screening tools is the lack of adequate training for healthcare providers. Many providers are not sufficiently trained in administering and interpreting these tools, which can lead to inconsistent application and variability in results ^41^. The ACE and ADAS-Cog have shown variability in results due to inconsistent application by healthcare providers. Without specialized training, healthcare providers may not administer these tools correctly, affecting the accuracy of the results. Training modules focusing on the proper administration and interpretation of tools should be made mandatory for healthcare providers.

Without a solid understanding of the cognitive assessment process, healthcare providers may also struggle to select appropriate tools, interpret results accurately, and communicate findings to patients and families. This gap in training not only affects the reliability of the assessments but also hampers the ability to engage patients effectively, which is essential for comprehensive cognitive evaluations.

### 5.6. Cultural Appropriateness

Finally, the cultural appropriateness of cognitive screening tools is a significant concern. Many currently used tools do not account for the diverse cultural and linguistic backgrounds of the Indian population ^42^. The MoCA, Demtec, MMSE, and CAMCOG were originally developed outside India, limiting their relevance to Indian cultural norms and practices. For example, Western cognitive frameworks may not align with how memory and cognition are understood and expressed in various Indian communities. Tools must be refined to reflect local cultural values and cognitive styles to ensure their effectiveness in India’s diverse populations. The lack of culturally sensitive assessments can lead to misunderstandings during evaluations and fail to accurately capture the cognitive status of individuals from diverse backgrounds ^43^.

Hence, the barriers to implementing cognitive screening tools in PHC settings are multifaceted and interrelated. Addressing these challenges requires a concerted effort to enhance the educational materials used in screenings, develop linguistically and culturally appropriate tools, invest in training for healthcare providers, and allocate resources effectively within PHC centres. Overcoming these barriers is essential for ensuring that cognitive screening tools are utilized effectively, ultimately leading to improved cognitive health outcomes for patients across diverse populations.

These results and analysis show that the existing cognitive screening tools in Primary Health Care (PHC) settings have significant gaps in their effectiveness, particularly concerning the educational and linguistic diversity of India’s population. While various screening tools are currently in use, none fully meet the needs of individuals with varying literacy levels and linguistic backgrounds. This inadequacy highlights an urgent need for culturally appropriate tools that are easy to administer and understand, especially in busy PHC environments where time and resources are limited.

Current tools also often fail to accommodate the realities of low literacy levels among patients, which can lead to misinterpretations and inaccurate assessments of cognitive function. Therefore, there is a pressing demand for the development and implementation of new screening instruments that cater specifically to these challenges. Such tools should prioritize simplicity in language and design, ensuring accessibility for all patients, regardless of their educational background.

Moreover, there is a critical need for cognitive screening tools that require minimal training for healthcare providers to administer effectively. Tools should be quick to use, allowing healthcare professionals to integrate them seamlessly into their existing workflows without extensive training. This approach will help mitigate educational bias, ensuring that patients from various educational backgrounds receive equitable assessments.

Additionally, the findings emphasize the importance of cultural sensitivity in tool development. Many existing tools do not account for the diverse cultural and linguistic backgrounds of the Indian population, which impacts their relevance and applicability. Tools designed with these factors in mind can enhance understanding and engagement during assessments.

Investing in training for healthcare providers to adopt and implement tools will also be essential. Ultimately, creating simple, quick, and culturally appropriate tools with minimal training requirements will significantly enhance the quality of cognitive assessments and improve care for patients in PHC settings.

## 6. Discussion

The scoping review highlights significant barriers to the effective use of cognitive screening tools in primary healthcare (PHC) settings in India, where diverse educational and linguistic backgrounds of patients pose unique challenges. Tools like the Mini-Mental State Examination (MMSE) and the Montreal Cognitive Assessment (MoCA) are commonly employed; however, their effectiveness is compromised due to these barriers. Research indicates that patients with lower educational attainment often struggle with the comprehension of tasks within these screening tools, leading to assessments that do not accurately reflect their cognitive abilities. This discrepancy can lead to misdiagnosis or delayed intervention, underscoring the importance of culturally and educationally sensitive approaches.

Language barriers further exacerbate the challenges faced in cognitive assessments. Many cognitive screening tools lack appropriate translations or adaptations for various Indian languages, leading to misunderstandings and inaccurate responses ^44,45^. The absence of linguistically tailored versions can result in false negatives, wherein a patient who may have cognitive impairment is incorrectly assessed as healthy, or false positives, wherein a cognitively healthy individual is misdiagnosed. Such inaccuracies not only affect individual patient care but also contribute to a broader public health challenge by potentially skewing dementia prevalence data and affecting resource allocation for care.

Training of healthcare providers is another critical area identified in this review. There is often insufficient training on how to administer and interpret these cognitive screening tools effectively ^46^. This lack of training can result in inconsistent administration and interpretation, further complicating the already challenging landscape of cognitive assessments in PHC settings. Studies have shown that provider training significantly enhances the reliability of cognitive assessments, suggesting that improving training programs for healthcare professionals is essential ^47^.

Cultural appropriateness also emerged as a vital concern. Many widely used cognitive screening tools do not adequately reflect the cultural nuances and context of the Indian population, impacting their relevance and applicability ^48^. Tools must not only be linguistically and educationally appropriate but also culturally sensitive to ensure they resonate with the diverse backgrounds of the population they aim to serve.

To address these gaps, there is an urgent need for the development of simple, quick, and less training-intensive cognitive screening tools that can be efficiently administered in busy PHC environments. Tools should be designed to accommodate individuals with low literacy levels, ensuring they can engage meaningfully with the assessment process. Moreover, incorporating input from local communities during the tool development phase can enhance cultural relevance and acceptance, facilitating a more accurate assessment of cognitive status across diverse populations ^49^.

In summary, addressing the barriers related to educational and linguistic diversity is crucial for enhancing the effectiveness of cognitive screening tools in India. By developing culturally appropriate and linguistically tailored tools and improving training for healthcare providers, we can improve the early detection and management of dementia. This not only enhances individual patient outcomes but also contributes to a more robust public health response to dementia in India. Future research should prioritize these areas to ensure equitable healthcare access for all segments of the population.

## 7. Strengths and Limitations of the Study

The primary strength of this scoping review lies in its comprehensive examination of cognitive screening tools specifically designed for dementia detection within primary healthcare settings in India. By focusing on tools that are culturally relevant and contextually applicable, this review highlights the importance of integrating local practices and needs into the evaluation of cognitive assessment methods. Additionally, the inclusion of diverse study designs and methodologies provides a well-rounded perspective on the effectiveness and applicability of these tools in real-world settings.

However, there are notable limitations to this review. Firstly, the scope was restricted to studies published in English, which may have excluded valuable insights from non-English literature that could enhance our understanding of cognitive screening practices in India. Furthermore, while we aimed to cover a broad range of cognitive assessment tools, the review may not fully encapsulate all existing tools and innovations in this rapidly evolving field. The focus on primary healthcare centres also limits the exploration of tools used in specialized settings, which might provide a different perspective on cognitive assessment. Lastly, while the review identifies several tools and their effectiveness, it does not delve deeply into the specific challenges faced by healthcare professionals in implementing these tools in everyday practice, such as training needs and resource allocation.

## 8. Conclusion

Cognitive screening tools play a vital role in the early detection and management of dementia within primary healthcare settings in India. However, the findings from this review underscore a critical need for tools that are culturally and linguistically appropriate, accommodating the diverse educational backgrounds of the Indian population. Existing tools like MMSE and MoCA, while widely used, do not fully meet these needs and face significant barriers that hinder their effectiveness.

To enhance the accuracy and reliability of cognitive assessments, it is essential to develop new tools that are easy to administer, require minimal training, and are sensitive to the linguistic and cultural diversity of the patient population. Furthermore, healthcare providers must receive adequate training to effectively utilize these tools in their practice. By addressing these gaps, we can improve the detection of cognitive impairments, leading to timely interventions that ultimately enhance the quality of care for individuals experiencing dementia. Future research should focus on validating these new tools and strategies to overcome implementation barriers, paving the way for more effective cognitive screening in India’s diverse healthcare landscape.

## Data Availability

All data produced in the present work are contained in the manuscript

